# Bacterial contamination of Healthcare workers’ mobile phones in Africa: a systematic review and meta-analysis

**DOI:** 10.1101/2022.05.27.22275700

**Authors:** Demisu Zenbaba, Biniyam Sahiledengle, Girma Beressa, Fikreab Desta, Zinash Teferu, Fikadu Nugusu, Daniel Atlaw, Zerihun Shiferaw, Ayele Mamo, Wogane Negash, Getahun Negash, Mohammedaman Mama, Eshetu Nigussie, Vijay Kumar Chattu

## Abstract

**Background:** Mobile phones are potential reservoirs for pathogens and sources of healthcare-associated illnesses. More microbes can be found on a mobile phone than on a man’s lavatory seat, the sole of a shoe, or a door handle. When examining patients, frequent handling of mobile phones can spread bacteria and provide a suitable breeding environment for numerous microorganisms. Nevertheless, evidence of bacterial contamination of mobile phones among healthcare workers in Africa was not conclusive. Thus, this meta-analysis and systematic review was conducted to estimate the pooled prevalence of bacterial contamination of mobile phones used by healthcare workers and the most frequent bacterial isolates in Africa.

**Methods:** We systematically retrieved relevant studies using PubMed/MEDLINE, Scopus, POPLINE, HINARI, Science Direct, Cochrane Library databases, and Google Scholar from 2009 to 2021 publication year. We included observational studies that reported the prevalence of bacterial contamination of mobile phones among healthcare workers. Two independent authors assessed the quality of the studies. The DerSimonian-random Laird’s effect model was used to calculate effect estimates for the pooled prevalence of bacterial contamination in mobile phones, as well as a 95% confidence interval (CI).

**Results:** Among 3882 retrieved studies, 23 eligible articles with a total sample size of 2,623 study participants were included in the meta-analysis. The pooled prevalence of mobile phones bacterial contamination among healthcare workers was 83.9% (95% CI: 80.6, 87.2%; I^2^ = 98%, p-value < 0.001). The most dominant type of bacteria isolated in this review was *coagulase-negative staphylococci* (CONS) which accounted for 44.5% of the pooled contamination rate of mobile phones used by healthcare workers, followed by *Staphylococcus aureus* (32.3%), and *Escherichia coli* (8.4%).

**Conclusion:** The review indicated that the contamination with a different bacterial isolate of mobile phones used by health care workers was high. The most dominant bacterial isolates were *Coagulase-negative staphylococci, Staphylococcus aureus*, and *Escherichia coli*. Hence, these findings would have implications for policymakers and resource allocation for preventive measures initiatives.

## Introduction

Currently, mobile phones have become essential accessories for healthcare workers and social life [1, 2]. Mobile phones have become an important part of the healthcare delivery system because they improve the quality of care and communication [1, 3]. It also makes interdepartmental communication easier, allowing for faster interactions within healthcare institutions and more efficient access to information for patient care [4, 5]. Despite all of the potential benefits, mobile phones play a critical role in becoming potential germ reservoirs and are known to induce healthcare-associated diseases [6-9].

A variety of bacteria, including skin flora and pathogenic bacteria, have been identified on the surface of mobile phones [10, 11]. In HICs, bacterial infections on healthcare personnel’s mobile phones range from 75 to 96% [12-19]. *Coagulase-negative staphylococci (CoNS)* and *Micrococcus* were the most commonly recovered bacteria, followed by *methicillin-sensitive and methicillin-resistant Staphylococcus aureus* (*MRSA*), *Acinetobacter*, and *Pseudomonas species* [12-19]. In low-and middle-income countries’ healthcare settings, bacterial contamination rates of mobile phones used by healthcare personnel ranged from 42% to 100%. The most prevalent bacteria isolated were *coagulase-negative staphylococci, Escherichia coli, Acinetobacter species, Pseudomonas species*, and *MRSA* bacteria [20-26]. Several infectious illnesses, including diarrhea, food poisoning, and wound infections, are caused by these bacteria [3, 27, 28].

The global burden of healthcare-associated infections (HAIs) is increasing, resulting in increased morbidity and death among patients and significant challenges for the healthcare system [7, 29, 30]. The cumulative incidence of HAIs ranges from 5.7 to 48.5% within African countries [31]. Contamination of inanimate gadgets used by healthcare professionals, such as cell phones, is one of the many sources of healthcare-acquired infections [30, 32]. More bacteria can be found on a mobile phone than on a man’s lavatory seat, the sole of a shoe, or a door handle [31, 33, 34]. Drug-resistant organisms such as *MRSA* and *vancomycin-resistant enterococci* (*VRE*) have also been found in hospital settings using mobile phones. The drug-resistant bacterium that can cause HAIs and is a public health issue was found to be responsible for 40% to 70% of the contamination of healthcare professionals’ mobile phones [14, 33].

When examining patients, frequent handling of mobile phones can spread bacteria and provide a suitable breeding environment for numerous microorganisms [10, 35, 36]. Although there has been some small-scale research on the bacterial contamination of mobile phones among healthcare workers, a comprehensive review and meta-analysis have not been conducted for Africa. Therefore, this systematic review and meta-analysis aimed to estimate the pooled prevalence of bacterial contamination of mobile phones used by healthcare workers and the most common bacterial isolates in Africa. Besides, we anticipated descriptively summarizing bacterial isolates’ antimicrobial susceptibility and multidrug resistance patterns.

## Methods

### Registration and protocol

This systematic review and meta-analysis (SRMA) was conducted to estimate the pooled prevalence of bacterial contamination of mobile phones among HCWs in Africa. To ensure the usefulness of this SRMA to the readers, we developed a transparent, complete, and accurate report of the purpose of this review, using the Preferred Reporting Items for Systematic Reviews and Meta-Analysis (PRISMA) criteria (Additional file 1 Table). This systematic review was carried out following the Joanna Briggs Institute (JBI) methodology for systematic reviews of a proportion of evidence [37]. The systematic review and meta-analysis were prospectively registered in PROSPERO (record ID: CRD42022306250, February 22, 2022).

### Search strategy

We systematically retrieved relevant studies using PubMed/MEDLINE, Scopus, POPLINE, HINARI, Science Direct, Cochrane Library databases, and Google Scholar from inception to March 25, 2022. All the databases were comprehensively searched to find potentially relevant papers published and unpublished between 2009 and 2021. All searches were limited to English-language papers. In addition to the electronic database search, Google was used to look for grey literature. We also looked for related studies in the reference lists of included studies. For the PubMed/MEDLINE search, the following phrases and keywords were used:[“Bacterial Contamination” OR “microbial contamination” OR “Contamination, equipment” AND “Cell Phones” OR “Mobile Phone” OR “Mobile Phones” OR “Smart Phones” OR “cellular Phones” AND Health Personnel” OR “HealthCare Providers” OR “Health Care Provider” OR “Provider, Health Care” OR “Healthcare Providers” OR “Healthcare Provider” OR “Provider, Healthcare” OR “Healthcare Workers” OR “Healthcare Worker” OR “Health Care Professionals” OR “Health Care Professional” OR “Professional, Health Care” AND Africa). We used database-specific subject headings linked with the above terms and keywords used in PubMed for the other electronic databases (Additional file 2).

### Eligibility criteria

#### Inclusion Criteria

The review process included all studies that met the following criteria: (1) studies that reported the magnitude of bacterial contamination from healthcare workers’ mobile phones surfaces, (2) studies published in English but conducted only in Africa at any given time, and (3) studies conducted using standard bacteriological techniques (i.e., swab method or settle plate sampling method) [32, 38, 39]. (4) Studies that accurately reported the swab culture growth rate for bacterial isolates, (5) all relevant free-of-charge full-text original research articles, and (6) all observational study designs, including published and unpublished studies, were all taken into account.

#### Exclusion Criteria

The study was excluded for the following reasons: inaccessible or irretrievable full-text articles after contacting the corresponding authors via email at least two times; reviews, commentaries, letters to the editor, conference proceedings, and abstracts; studies with unclear methods; reports from inanimate objects other than mobile phones (such as Stethoscopes, BP apparatus, and patient beds); studies conducted on non-healthcare workers; and studies that did not report the outcome of interest.

### Assessment of outcome variables

The primary outcome variable was the prevalence of bacterial contamination of mobile phones used by healthcare workers, as defined by the included studies’ operational definition. The prevalence of mobile phone bacterial contamination was calculated by dividing the total number of swabs collected from the mobile phones of healthcare professionals by the total number of swabs taken from mobile phones of healthcare workers and multiplying by 100. The second objective of this study was to descriptively characterize the most common types of bacteria isolated from healthcare workers’ mobile phones and their drug sensitivity and resistance patterns, utilizing studies that were included.

### Operational definitions

#### Non-selective bacteria isolation method

Culture mediums such as blood agar and nutrient agar can grow a wide variety of bacteria [39].

#### Selective bacteria isolation method

A culture medium such as MacConkey agar is more selective to isolate ‘bile tolerant’ bacteria found in the large intestine [39].

### Study selection and data extraction

All the retrieved citations were imported into EndNote version X8 and duplicates were removed. The JBI data extraction format was used to extract the data [40]. Based on the established inclusion criteria, two authors (DZ and BS) independently assessed and identified papers by their titles, abstracts, and full texts. Any disagreements that arose were resolved by consensus or with the additional author/s (DZ and GB). The data extraction format included the primary author, publication year, country, study area, bacteria isolation method, optimum temperature, incubation period, the most prevalent types of bacteria isolated, isolated bacteria drug sensitivity, isolated bacteria drug resistance, sample size, and prevalence of mobile phone bacterial contamination.

### Assessment of risk of bias

The quality of the appended studies was assessed using the JBI meta-analysis of statistics assessment and review instrument (MAStARI) quality rating tool [40, 41]. An appropriate sampling frame, proper sampling technique, study subject and setting description, sufficient data analysis, use of valid methods for the identified conditions, a valid measurement for all participants, using appropriate statistical analysis, in a valid and reliable outcome measure, with a 50% or higher overall score considered low risk of bias, are among the JBI parameters. As a result, bias risks were classified as low (total score of 2), moderate (total score of 3-4), or high (total score of > 5) [41]. Two independent authors rated the quality of the included studies (DZ and BS). Any disagreements that arose were addressed through consensus. Finally, papers with a score of 5 or higher were ruled out as having a significant risk of bias (Additional file 3).

### Data synthesis

Before being evaluated, the data was extracted into a Microsoft Excel file. The data were analyzed using STATA software version 16. The standard errors of the included studies were determined using the formula (SE = p (1p)/n). To investigate heterogeneity in the stated proportion, the I^2^ statistics and p-values of the Cochrane Q-test were utilized. The Cochrane Q-test p-values are less than 0.1 and are deemed to indicate the presence of heterogeneity among studies. To assess the percentage of total variance owing to heterogeneity across trials, we used the Higgins I^2^ test statistics [41]. Although no specific criterion exists for when heterogeneity becomes substantial, some researchers suggest low heterogeneity when I^2^ values are between (25–50%), moderate (50–75%), and high (> 75%) [41]. Because the test statistic revealed significant heterogeneity among the research (I^2^ = 98%, p-value 0.001), the DerSimonian-influence Laird’s was analyzed using a random-effects model. The effect sizes were calculated as a percentage with a 95% confidence interval (CI). There was a lot of variation in the included studies in this review according to the I^2^ category. We used subgroup analysis by sub-region, study area, bacteria isolation method, sample size, and publication year to find the source of variation. The meta-analysis findings were displayed using a forest plot. A funnel plot was employed in conjunction with meta-regression to assess publication bias. The plot resembles an asymmetrical, huge, inverted funnel in the absence of publication bias. Egger’s weighted regression and Begg’s rank correlation tests (p-value < 0.05) were used to objectively assess publication bias; however, only Egger’s test was shown to be statistically significant (p-value = 0.001). To test the findings’ robustness, a leave-one-out sensitivity meta-analysis was used.

## Results

A total of 3,882 articles were identified after a thorough search of the literature. Of these articles, 2,951 duplicates were removed, and 931 were screened only based on their titles and abstracts. Following the exclusion of 844 articles, a total of 87 full-text papers were verified for eligibility using the pre-determined criteria, with 64 articles being excluded. Finally, 23 articles [21–25, 42-59] that satisfied the criteria were included in the meta-analysis (Fig. 1).

**Figure 1:**
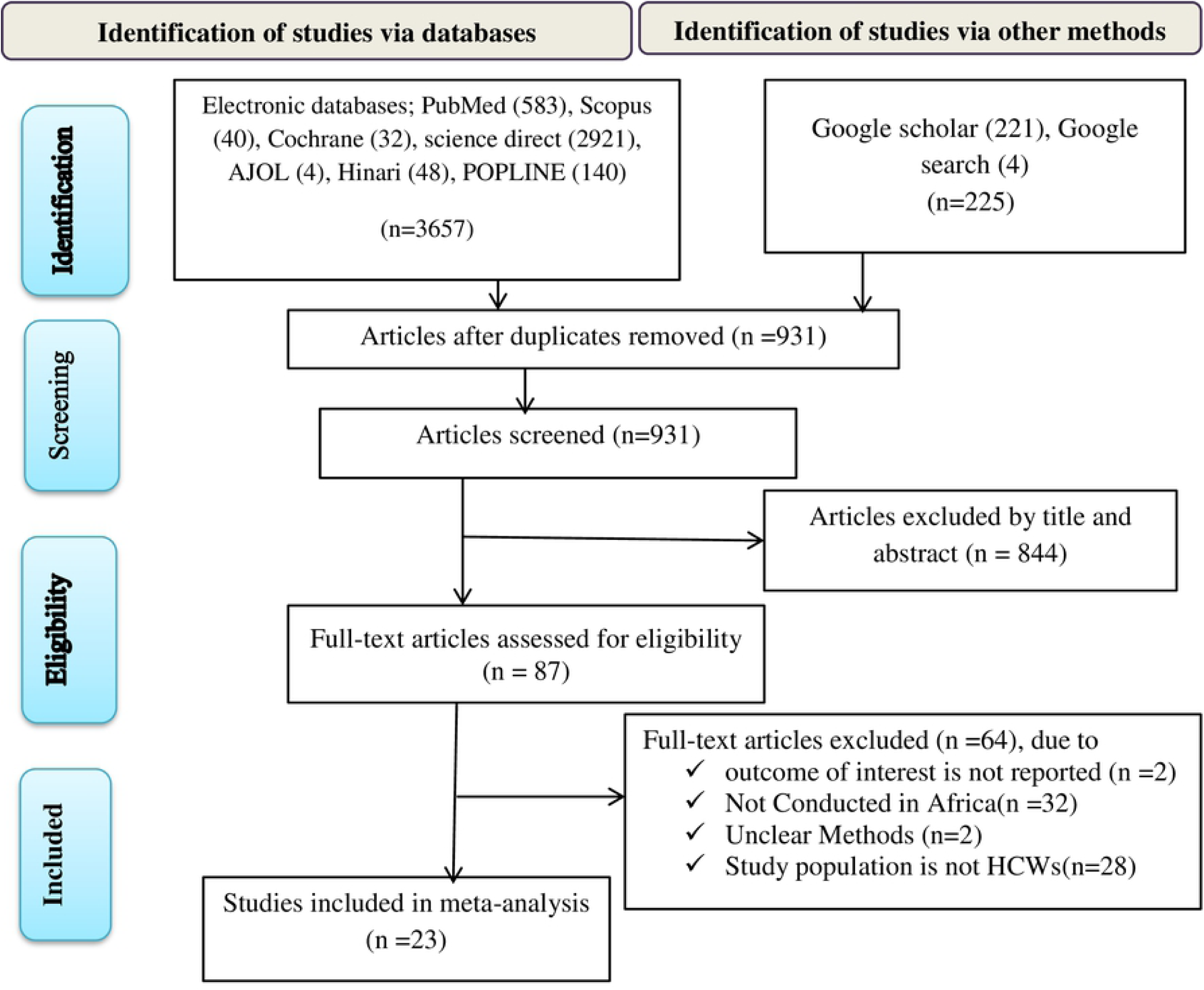
Flow chart of study selection for systematic review and meta-analysis of bacterial contamination of mobile phones among healthcare workers in Africa.

### Descriptions of the included studies

All included studies were cross-sectional by design and were published between 2009 and 2021. The current meta-analysis used 2,623 mobile phones from healthcare professionals to estimate the pooled proportion of bacterial contamination. In terms of sub-regional distribution, nine studies were from Eastern Africa [22, 24, 25, 42, 44–46, 58], four studies from Western [21, 51, 52, 55], seven studies from Northern [23, 47–50, 54, 57], one study from Southern [56], and two studies from middle African countries [53, 59] (Table 1).

**Table 1:**
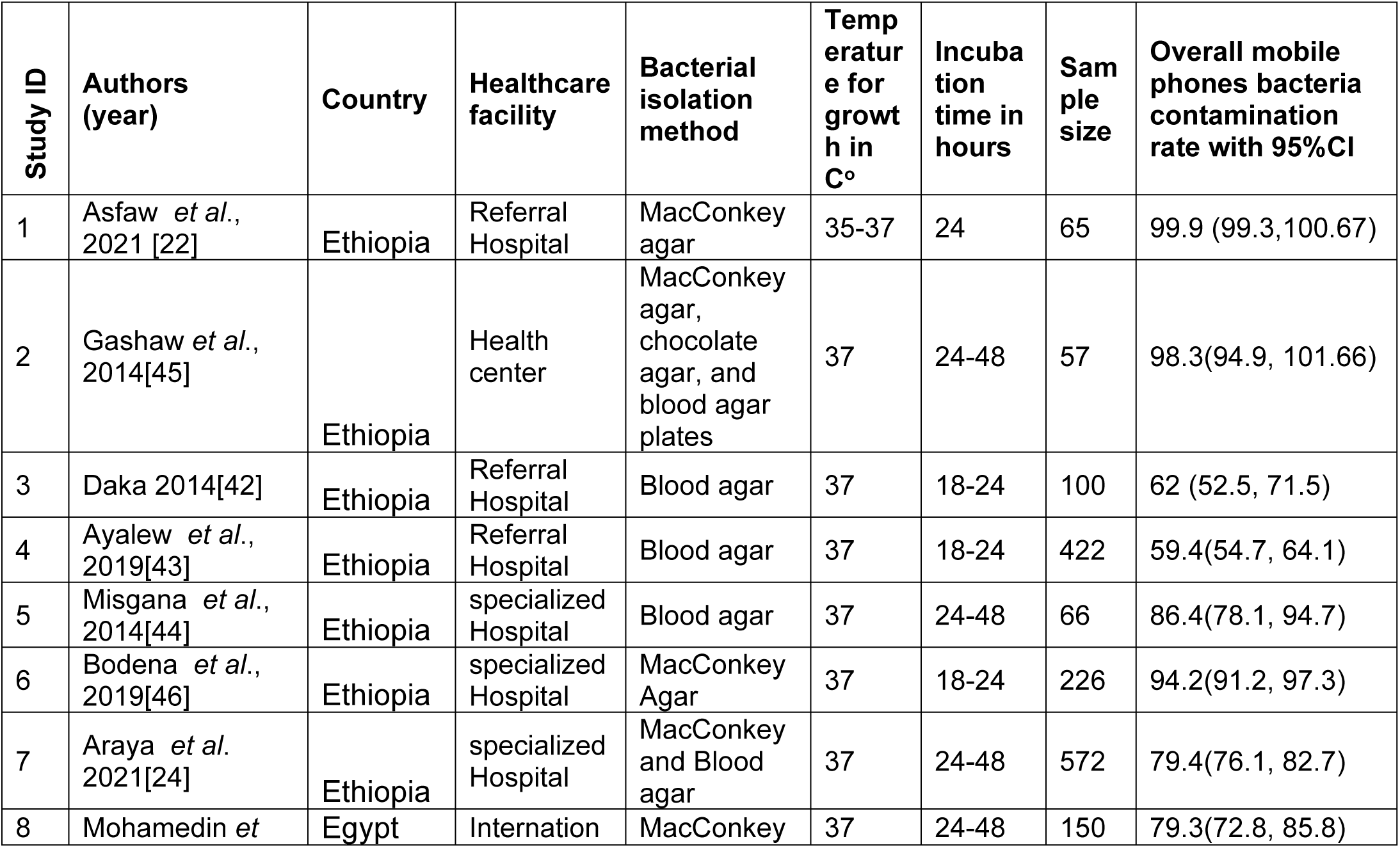
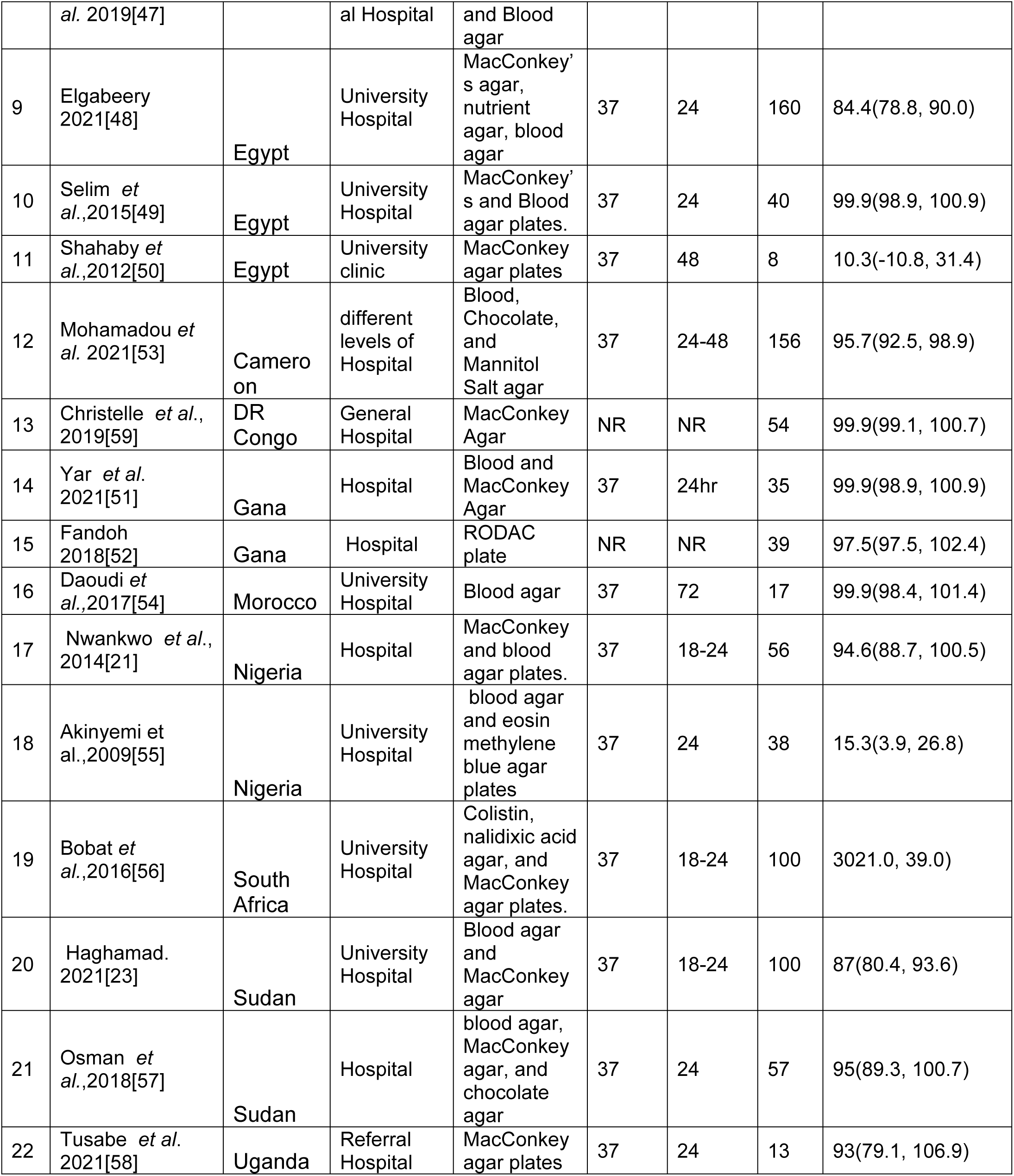
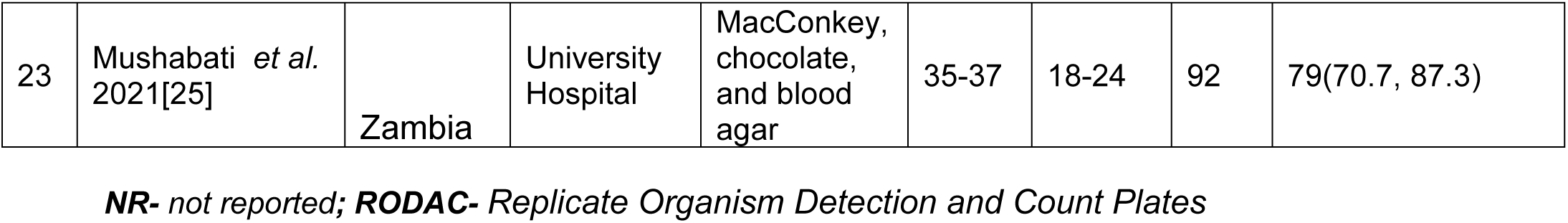
Descriptive summary of 23 studies included in the meta-analysis to estimate pooled prevalence of HCWs bacterial contamination of mobile phones in Africa.

### Prevalence and types of bacterial isolates

The pooled prevalence of bacterial contamination of mobile phones used by healthcare professionals in Africa was 83.9%; 95% CI: (80.6, 87.2%) (Fig.2). the high heterogeneity was showed among included studies (I^2^ = 98%, p = 0.001). As a result, a random effect model was used to estimate the pooled prevalence of bacterial contamination of mobile phones of healthcare personnel. A univariate meta-regression analysis was performed using variables such as year of publication, quality score, and sample size to identify credible sources of heterogeneity. Accordingly, the year of publication was a significant source of variability among the variables included in the studies (Table 2).

**Figure 2:**
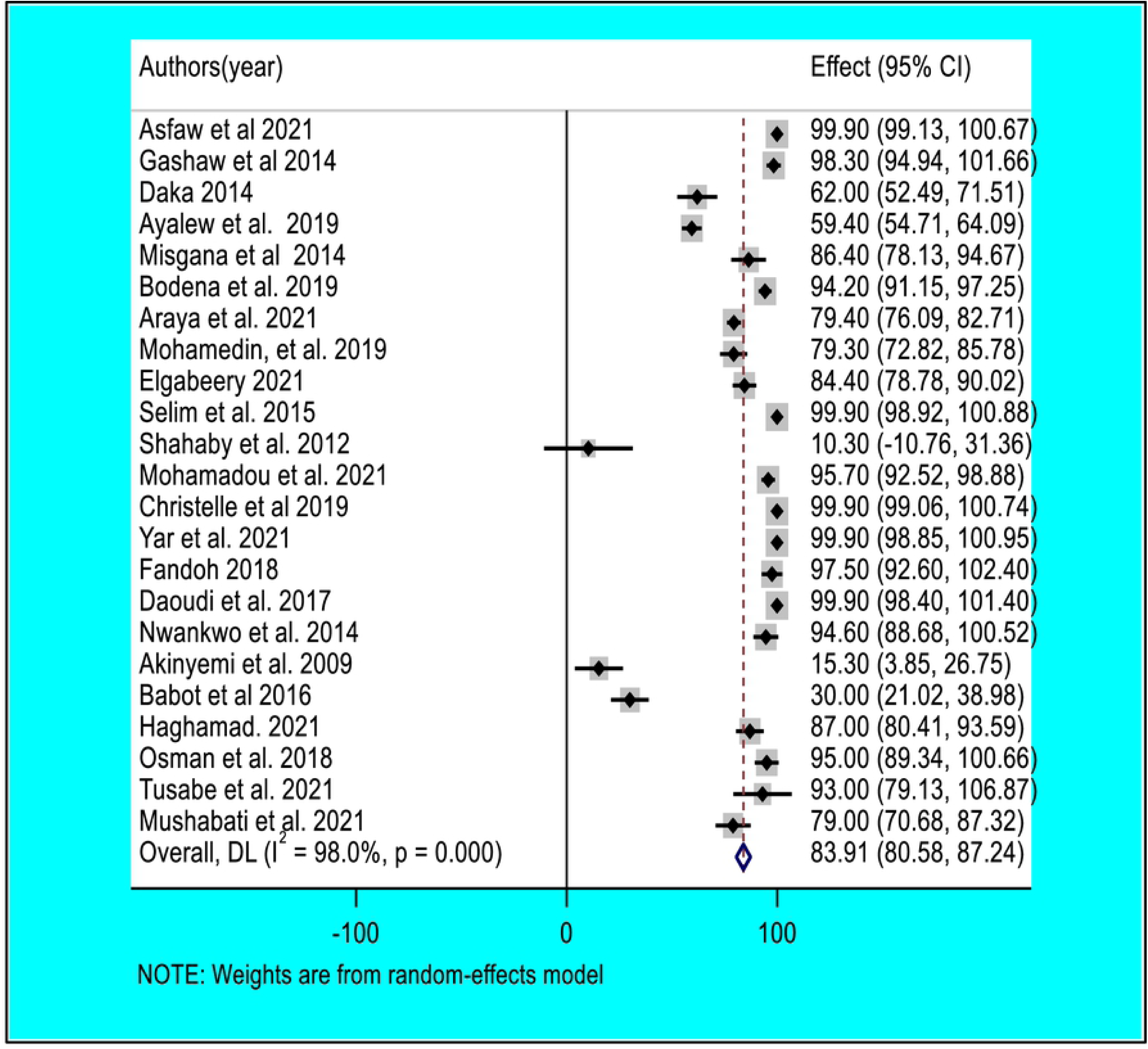
Forest plot of pooled bacterial contamination rate of mobile phones used by healthcare workers in Africa.

**Table 2:** Factors with the heterogeneity of HCWs mobile phones bacterial contamination based on univariate meta-regression.

| Variable | Coefficient | p-value | 95% CI |
| --- | --- | --- | --- |
| Year of publication | 3.42 | <0.001 | 2.28, 4.56 |
| Sample size | -0.03 | 0.003 | -0.05, 0.01 |
| Sub-region | 0.42 | 0.781 | -2.53, 3.37 |
| Culture media | -2.28 | 0.414 | -7.74, 3.19 |
| Quality score | 1.88 | 0.722 | -8.48, 12.24 |

The most prevalent bacteria in this review were *coagulase-negative staphylococci* (*CONS*), which accounted for 44.5% of the pooled contamination rate (95% CI: 34.3, 54.8%) of mobile phones used by healthcare workers, followed by *Staphylococcus aureus*, which accounted for 32.3% of the pooled contamination rate (95% CI: (34.3, 54.8%) of mobile phones used by healthcare workers (24.3, 40.2%). On the other hand, the gram-negative bacterium *Escherichia coli* was found in 8.4% of mobile phones used by healthcare personnel (95% CI: (5.1, 11.7%)) (Fig. 3-5).

**Figure 3:**
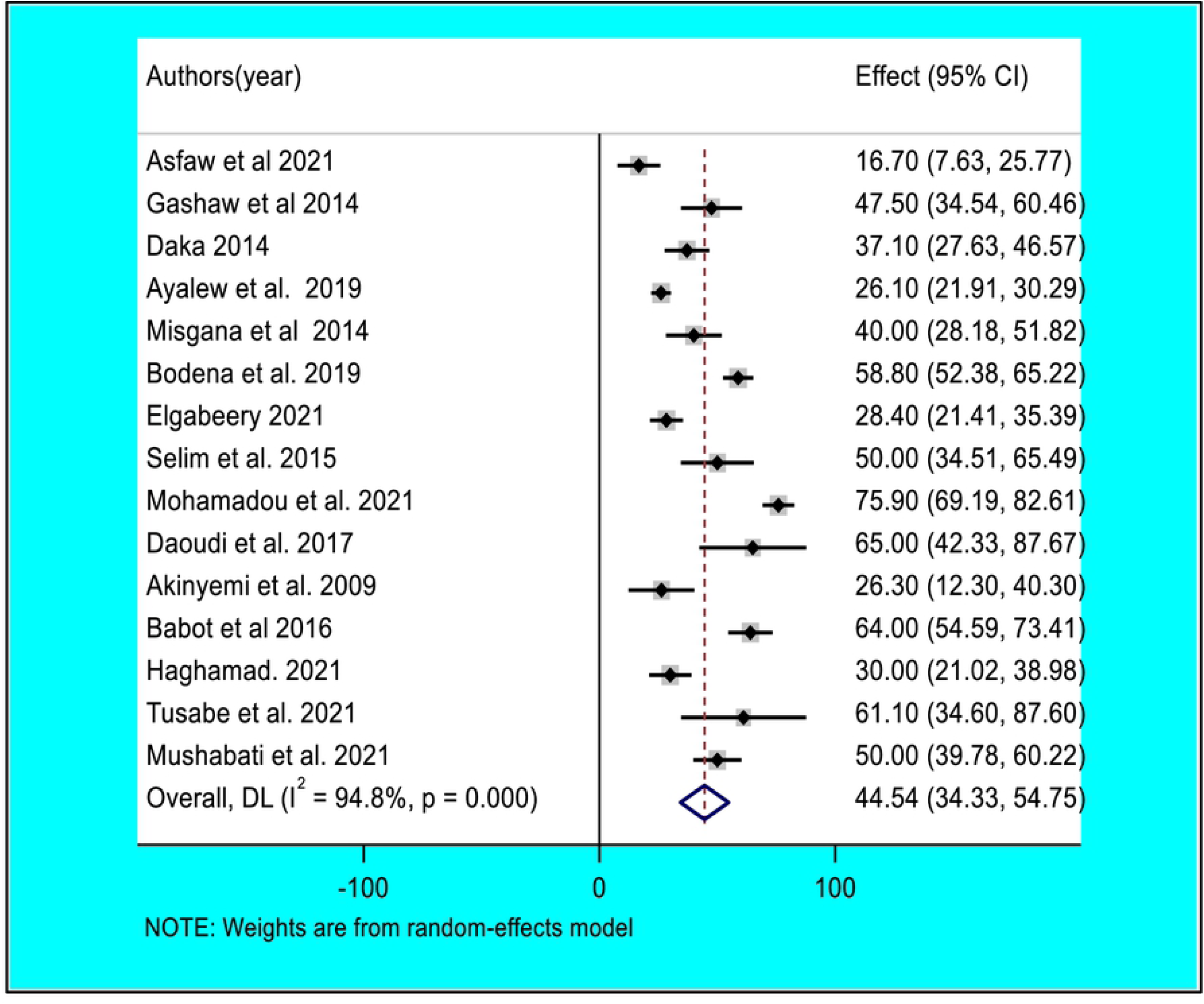
Forest plot of pooled contamination rate of mobile phones of healthcare workers by coagulase-negative staphylococci in Africa.

**Figure 4:**
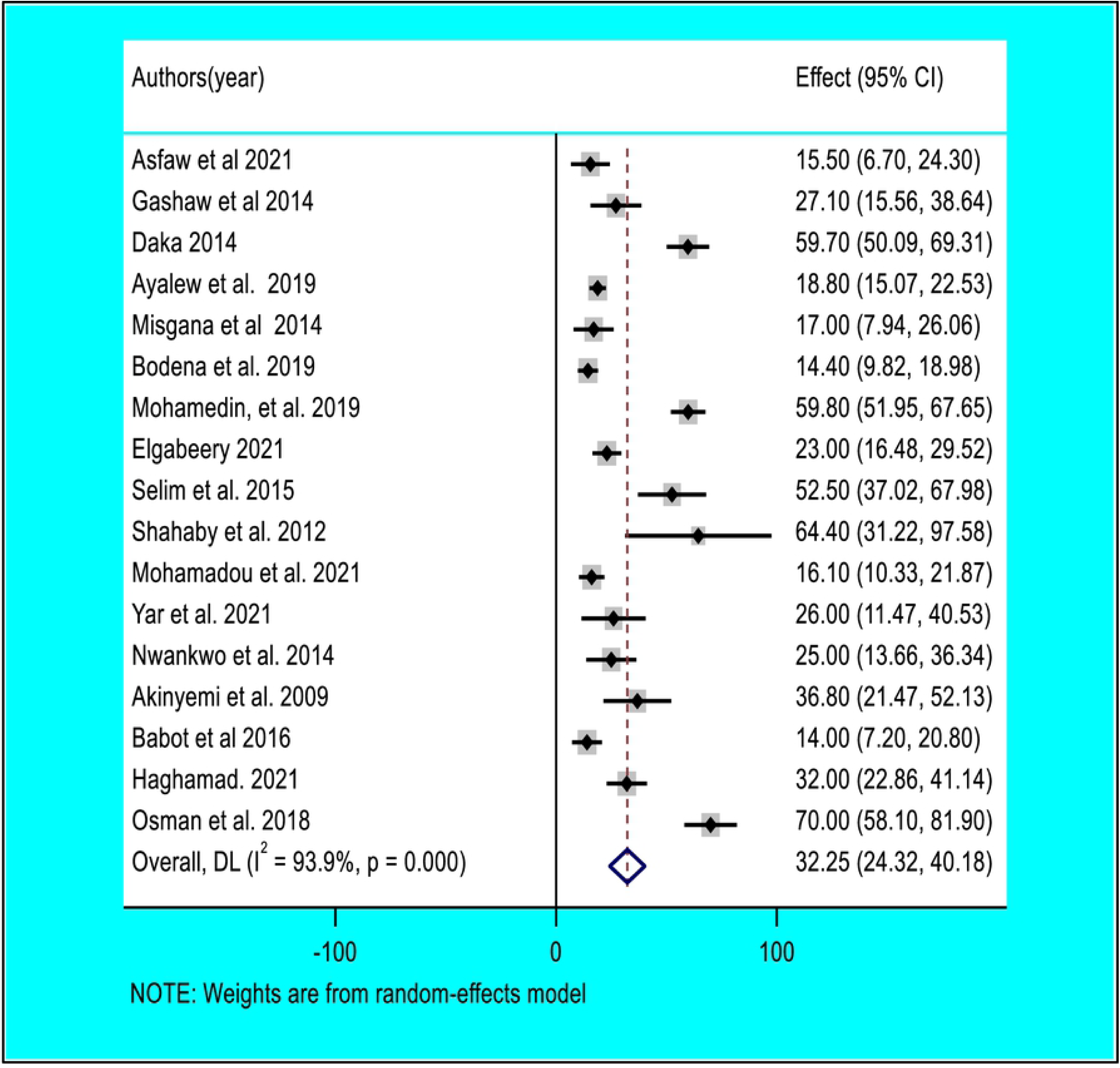
Forest plot of pooled contamination rate of mobile phones of healthcare workers by Staphylococcus aurous in Africa.

**Figure 5:**
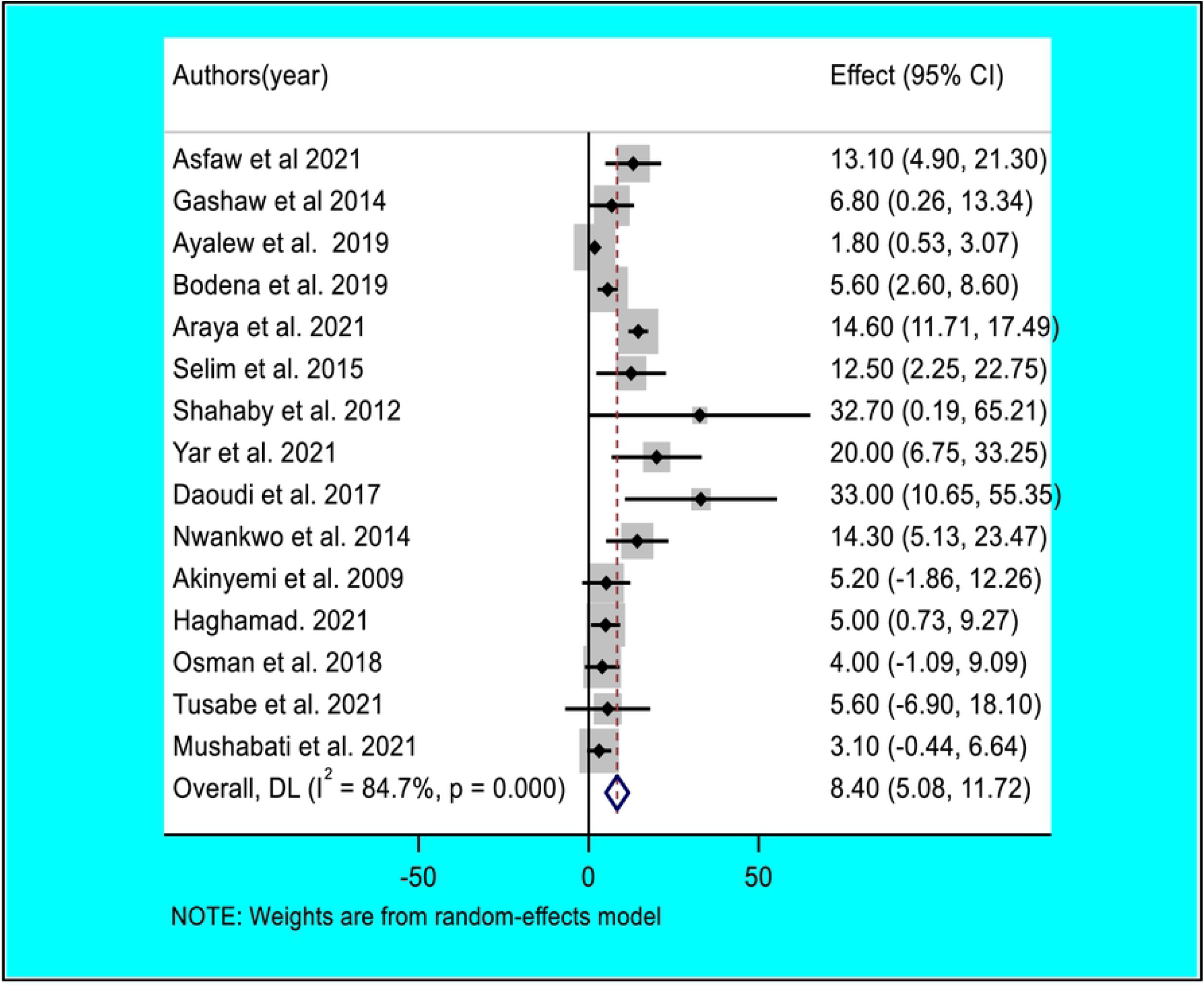
Forest plot of pooled contamination rate of mobile phones of healthcare workers by Escherichia Coli in Africa.

### Sensitivity analysis

The findings were put to the test using a leave-one-out sensitivity analysis. The random-effects model was shown to be robust, and according to the sensitivity analyses, no single study had an effect on the pooled rate of bacterial contamination of mobile phones used by healthcare personnel. The pooled mobile phone bacterial contamination was nearly equal to the real effect magnitude when a single study was eliminated from a meta-analysis (Fig. 6).

**Figure 6:**
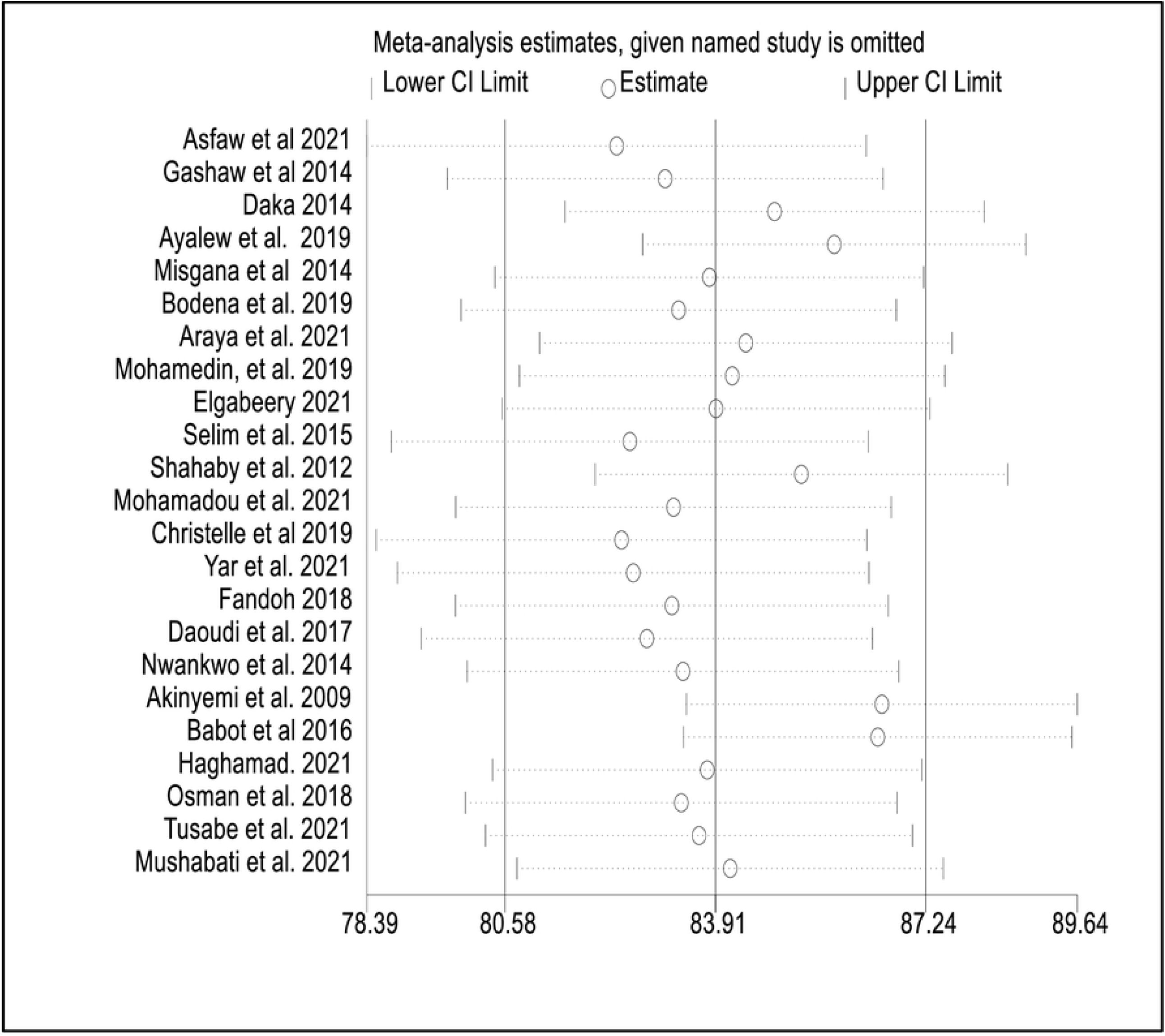
Sensitivity analysis of mobile phones bacterial contamination removed at a time: contamination rate and 95% confidence interval among healthcare workers in Africa.

### Publication bias

The funnel plot was used to examine the publication bias. The funnel plot demonstrated that the item distribution was consistent. We employed Begg’s and Egger’s tests to objectively confirm the asymmetry. In the prevalence of bacterial contamination of mobile phones used by healthcare workers, Egger’s test indicated evidence of publishing bias (p 0.001). However, Begg’s test revealed no indication of publication bias (p = 1.999). (Fig. 7). As a result, a non-parametric trim-and-fill analysis of the publication bias linear estimator was performed on the left, with the pooled prevalence of bacterial contamination set to 79.8% (Fig.8).

**Figure 7:**
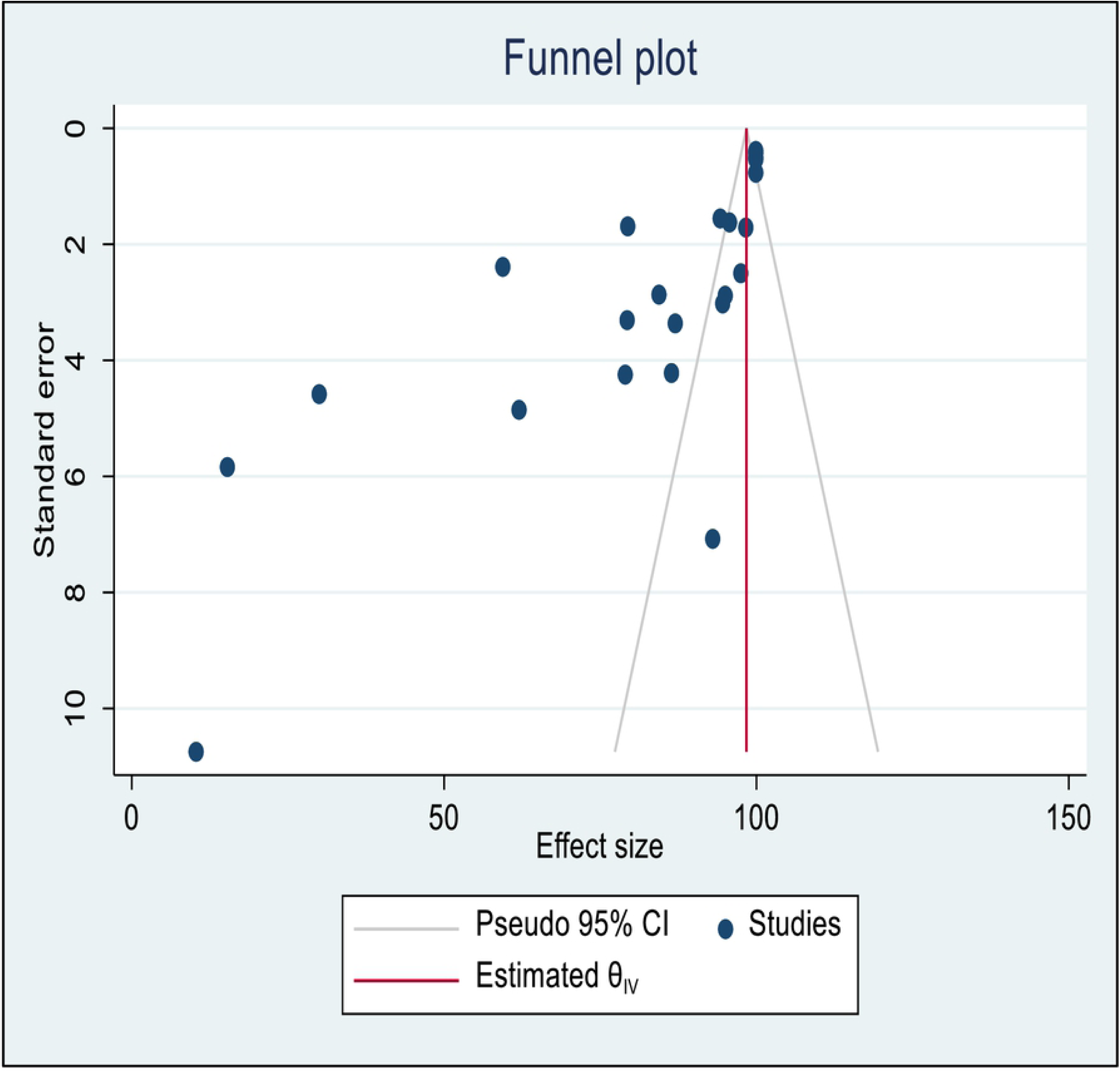
Funnel plot with 95% confidence limits of the pooled bacterial contamination rate of mobile phones used by healthcare workers in Africa.

**Figure 8:**
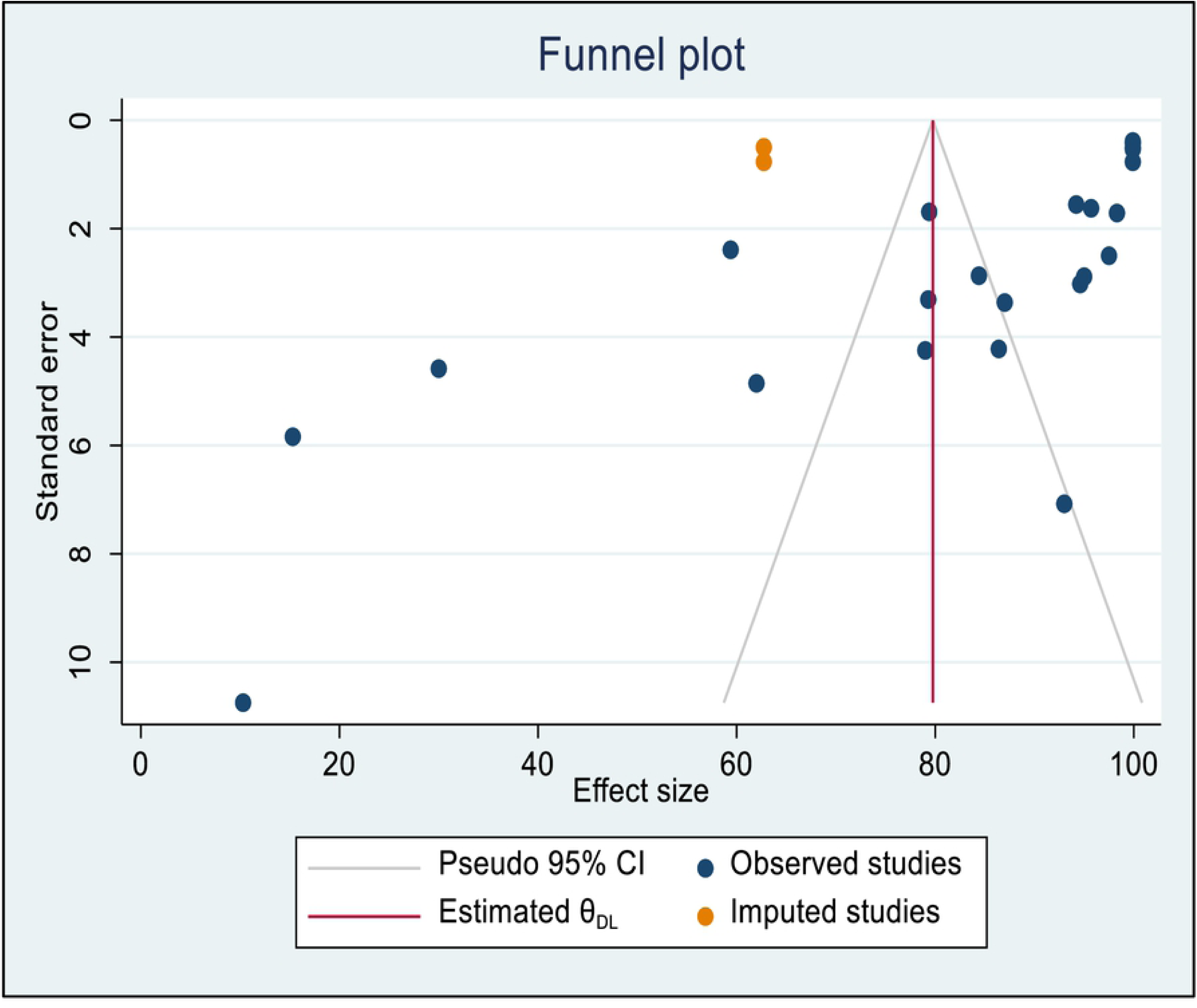
Funnel plot trim fill analysis with 95% confidence limits of the pooled bacterial contamination rate of mobile phones used by healthcare workers in Africa.

### Subgroup analysis

This meta-analysis used subgroup analysis based on the country’s sub-regions, study setting, and sample size. As a result, the northern African countries had the greatest pooled prevalence of bacterial contamination of mobile phones, at 87.3% (95% CI: (81.6, 93.0%), followed by the eastern African countries, at 83.62 %(95 % CI: 74.40, 92.84%). A subgroup analysis depending on the year of publication was also performed. The combined percentage of bacterial contamination in mobile phones among studies conducted from 2009 to 2014 and 2015 to 2021 was 62.5% and 88% respectively. The prevalence of bacterial contamination on mobile phones was 95.2% in studies that used a selective bacterial isolation method. However, it was found to be 70.4% in studies that used a non-selective bacterial isolation method, and 86.3% among studies that used both (selective + non-selective) bacterial isolation methods. A substantial source of variability was observed across the country’s sub-regions, year of publication, types of healthcare facilities, and bacterial isolation methods of included studies in all subgroup analyses (Table 3).

**Table 3:**
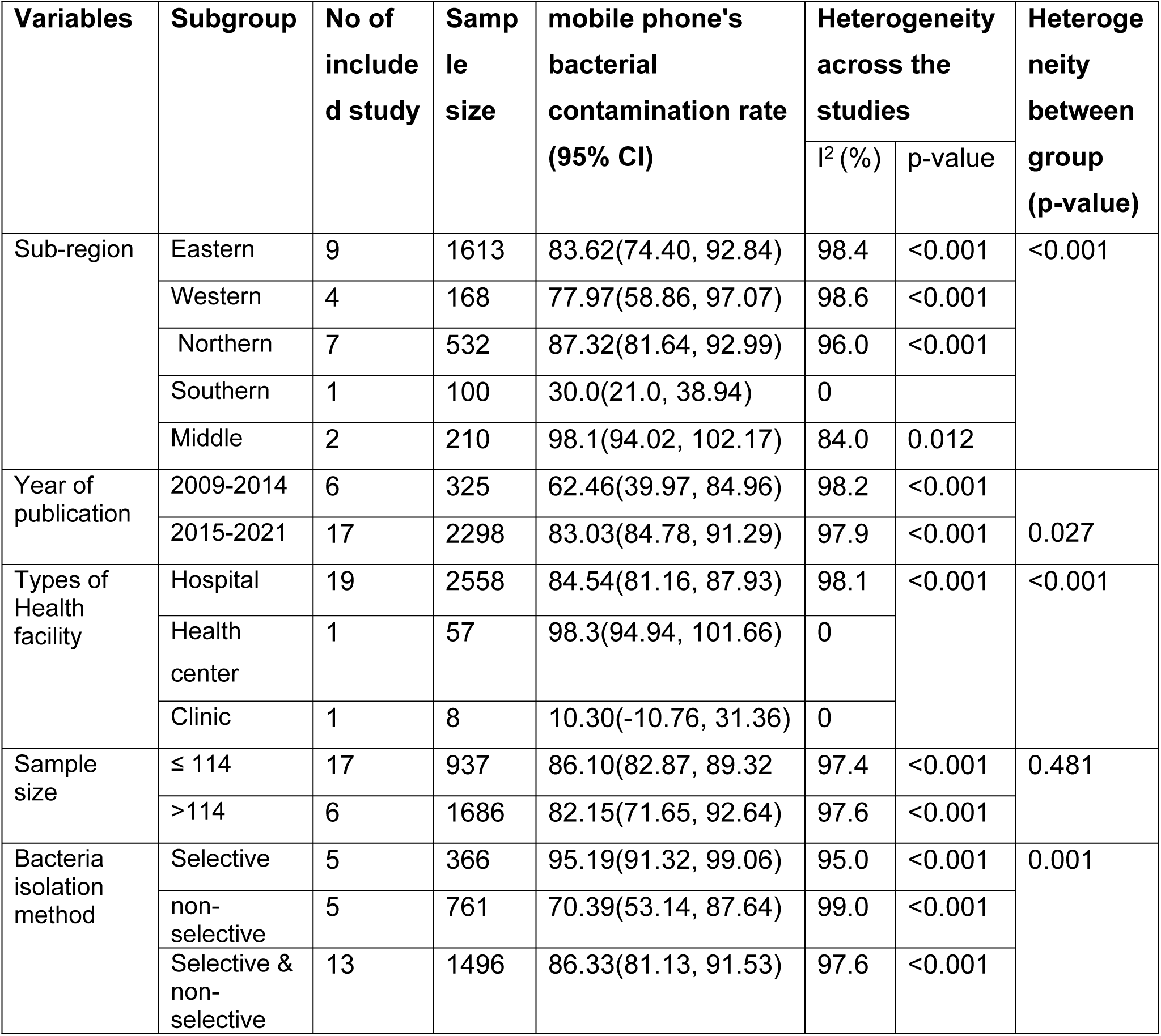
The subgroup rate of mobile phone bacterial contamination among healthcare workers in Africa (2009–2021)

| Variables | Subgroup | No of included study | Sample size | mobile phone's bacterial contamination rate (95% CI) | Heterogeneity across the studies |  | Heterogeneity between group (p-value) |
| --- | --- | --- | --- | --- | --- | --- | --- |
|  |  |  |  |  | I<sup>2</sup> (%) | p-value |  |
| Sub-region | Eastern | 9 | 1613 | 83.62(74.40, 92.84) | 98.4 | <0.001 | <0.001 |
|  | Western | 4 | 168 | 77.97(58.86, 97.07) | 98.6 | <0.001 |  |
|  | Northern | 7 | 532 | 87.32(81.64, 92.99) | 96.0 | <0.001 |  |
|  | Southern | 1 | 100 | 30.0(21.0, 38.94) | 0 |  |  |
|  | Middle | 2 | 210 | 98.1(94.02, 102.17) | 84.0 | 0.012 |  |
| Year of publication | 2009-2014 | 6 | 325 | 62.46(39.97, 84.96) | 98.2 | <0.001 | 0.027 |
|  | 2015-2021 | 17 | 2298 | 83.03(84.78, 91.29) | 97.9 | <0.001 |  |
| Types of Health facility | Hospital | 19 | 2558 | 84.54(81.16, 87.93) | 98.1 | <0.001 | <0.001 |
|  | Health center | 1 | 57 | 98.3(94.94, 101.66) | 0 |  |  |
|  | Clinic | 1 | 8 | 10.30(-10.76, 31.36) | 0 |  |  |
| Sample size | ≤ 114 | 17 | 937 | 86.10(82.87, 89.32) | 97.4 | <0.001 | 0.481 |
|  | >114 | 6 | 1686 | 82.15(71.65, 92.64) | 97.6 | <0.001 |  |
| Bacteria isolation method | Selective | 5 | 366 | 95.19(91.32, 99.06) | 95.0 | <0.001 | 0.001 |
|  | non-selective | 5 | 761 | 70.39(53.14, 87.64) | 99.0 | <0.001 |  |
|  | Selective & non-selective | 13 | 1496 | 86.33(81.13, 91.53) | 97.6 | <0.001 |  |

### Narrative review

#### Antimicrobial susceptibility and multidrug resistance patterns

We descriptively explained the antimicrobial susceptibility and multidrug resistance of bacterial isolates using 14 studies [21, 22, 24, 25, 42–46, 53–56, 58]. According to an Ethiopian study, bacterial isolates had a greater rate of resistance *to penicillin* (84%), *ampicillin* (81%), and *tetracycline* (81%). Nevertheless, a study conducted in Nigeria revealed that over 75% of bacterial isolates were sensitive to *fluoroquinolone* and *ceftriaxone* (Table 4).

**Table 4:** Summary of antimicrobial susceptibility and multidrug resistance pattern of bacterial isolates in Africa.

| Authors(Year) | Country | Antimicrobial susceptibility of the bacterial Isolates | MDR Pattern of Bacterial isolates |
| --- | --- | --- | --- |
| Asfaw *et al.* 2021 [22] | Ethiopia | Lower resistance rate against *ciprofloxacin* (24%), *gentamycin* (23%), and *nicol* (18 %). | -The overall MDR prevalence was found to be (42.9%).<br>-Bacterial isolates showed a higher resistance rate against *penicillin* (84%), *ampicillin* (81%), and *tetracycline* (42%). |
| Gashaw *et al.*, 2014[45] | Ethiopia | -*Ceftriaxone* and *ciprofloxacin* were effective against 71.7% and 89.1%, respectively, the Gram-positive bacteria<br>-About 87.5% of *S. aureus*, 89.3% of *CONS*, and all *S. pyogenes* isolate.<br>-Among the Gram-negative bacteria *E. coli* was found to be 100% sensitive to *ciprofloxacin*, *gentamycin*, and *trimethoprim-sulfamethoxazole*. | -More than half (52.2%) and 60.9% of Gram-positive bacteria were resistant to *amoxicillin* and *trimethoprim-sulfamethoxazole*.<br>-*E. cloacae* were 100% resistant to *ceftriaxone*, *ciprofloxacin*, *amoxicillin*, and *chloramphenicol* |
| Misgana *et al.*, 2014[44] | Ethiopia | -About 81% of the isolates tested for antimicrobial susceptibility | -About 39.40% of *S. aureus* isolates were *MARSA*, of which 38.50% (5/13) were *vancomycin-resistant*. |
| Bodena *et al.*, 2019[46] | Ethiopia | -*Ceftriaxone* (80.6%), *ciprofloxacin* (77.3%), and *gentamicin* (72.7%) showed higher activity against bacterial isolates,<br>-*Ampicillin* and *trimethoprim-sulfamethoxazole* had less effect, with a resistance rate of 61.6% and 56.9%, respectively. | - Overall prevalence of MDR bacterial isolates was 69.9%.<br>-Amongst all the bacterial isolates, *Pseudomonas sp.* (87.5%), *Klebsiella sp.* (86.7%), and *Citrobacter sp.* (75%) showed MDR |
| Araya *et al.* 2021[24] | Ethiopia | - High-level resistance to *ampicillin* was observed in 100% of all isolated species except for : *Citrobacter* (80%) and *K. pneumonia* (92%) *K. niae*, and *E. coli* also exhibited substantial resistance to *ampicillin* (92% and 100%, respectively) | -About 79.2% of the *ESBL*-producing isolates showed multidrug resistance.<br>-*K. oxytoca*, *Salmonella spp.*, *P. vulgaris*, and *P.mirabilis* showed 100% MDR characteristics |
| Mohamedin *et al.*, 2019[47] | Egypt | -About 100% of *S. aureus* was sensitive to Kanamycin and *Trimethoprim sulphamethoxazole* | -Around 98.2% of *S.aureus* was resistant to Methicillin, *Oxacillin*, and *Ampicillin* antibiotics |
| Mohamadou *et al.* 2021[53] | Cameroon | -*Ciprofloxacin* (85.7%), *Ofloxacin* (78.6%), *Gentamicin* (71.4%) to bacterial isolates | Not reported |
| Daoudi *et al.*, 2017[54] | Morocco | Not reported | -Of the 17 mobile phones, 6 were contaminated with multi-drug-resistant pathogens, with a contamination rate of 35%. |
| Nwankwo *et al.*, 2014[21] | Nigeria | Not reported | -High level of bacterial isolate resistance against *cotrimoxazole*, *tetracycline* and *ampicillin*, *gentamicin*, *ceftriaxone*, and *ciprofloxacin* |
| Akinyemi *et al.* 2009[55] | Nigeria | -Over 75% of the isolates were susceptible to the *fluoroquinolone* and *ceftriaxone* antibiotics. | Not reported |
| Bobat *et al.*, 2016[56] | South Africa | -All of the *S. aureus* isolated were *methicillin/cloxacillin* sensitive. | Not reported |
| Osman *et al.*, 2018[57] | Sudan | -*Staphylococcus aureus* isolates' sensitivity to *Oxacillin* was 1.40%. | -*Staphylococcus aureus* isolates were 98.6% resistant to *Oxacillin* |
| Tusabe *et al.* 2021[58] | Uganda | Not reported | -About 45% of the organisms were multidrug-resistant.<br>-Resistance was major to *penicillin*, *cotrimoxazole*, *ciprofloxacin*, and *Gentamycin* |
| Mushabati *et al.* 2021[25] | Zambia | -Most of the isolates were susceptible to first-line antimicrobial agents, except penicillin which showed 100% resistance for all Gram-positive isolates.<br>- *S. aureus* was susceptible to *ciprofloxacin* (88%), *clindamycin* (88%), *gentamicin* (84%), *cotrimoxazole* (50%) and *erythromycin* (50%) | Resistance to *cefoxitin* was detected in 25% (6/24) of *S. aureus* and 48% (21/49) of CoNS |

CONS: Coagulase-negative staphylococci; ESBL: Extended-spectrum beta-lactamase; MDR: Multidrug resistance

## Discussion

Mobile phones (MPs) are now widely used worldwide and are regarded as one of the essential gadgets. This technology is thought to be one of the most significant threats to human health [60, 61]. Healthcare workers’ (HCWs’) continuous handlings of MPs promote the spread of healthcare-associated illnesses [62]. In addition, pathogenic organisms colonizing mobile phones may increase antibiotic resistance [1]. The objective of this systematic review and meta-analysis was to estimate the pooled prevalence of bacterial contamination of mobile phones used by healthcare workers in Africa. As a result, 83.9% of mobile phones were found to be contaminated with bacteria. Mobile phone bacterial contamination is responsible for different infectious illnesses and increases the burden of nosocomial infections unless standard guidelines for using and cleaning mobile phones in health care settings are established [1, 3, 27, 28]. On the other hand, bacterial contamination of MPs could be a significant concern influencing the execution of efficient infection prevention measures, thus jeopardizing efforts to limit cross-contamination [63]. This review’s result was somewhat higher than that of a meta-analysis in Egypt, which reported a pooled prevalence of bacterial contamination of mobile phones of 78% [26]. Similarly, this review finding was consistent with findings from a systematic review published elsewhere [64]. The variation in bacterial contamination of mobile phones could be due to the varying levels of hand hygiene practiced by healthcare staff, the different models of mobile phones utilized, and bacterial isolation method [17, 65]. Furthermore, the type and load of bacterial contamination are known to be influenced by the design of touchscreen phones and the type of keypad surface [66-68].

We conducted a sub-group analysis based on country sub-region, finding that research from northern African countries had the highest incidence of bacterial contamination in mobile phones. In contrast, studies from southern African countries had the lowest prevalence. Compared to research conducted in other countries in sub-regions, most of the papers included in this review were from eastern and northern African countries, and differences in healthcare facilities could explain regional heterogeneity. Another reason for the disparities could be variances in healthcare personnel’s hand hygiene standards, mobile phone cleaning methods, and phone types (touchscreen versus Keypad surface). As a result of our findings, it may be necessary to encourage all African countries to achieve a zero prevalence of bacterial contamination in mobile phones.

A subgroup analysis was also performed using the year of publication and the method of bacterial isolation. As a result, studies conducted from 2015 to 2021 found a higher incidence of bacterial contamination in mobile phones than studies conducted from 2009 to 2014, which demonstrated a lower frequency of bacterial contamination. This disparity could be because smartphones or screen touch mobile phones, which have a high contamination rate and have been used by healthcare personnel in recent years, have a high contamination rate. In terms of bacterial isolation methods, research using selective bacterial isolation methods like MacConkey had the highest frequency of bacterial contamination on mobile phones compared to studies using non-selective and both (selective & non-selective) bacterial isolation methods. These differences could be related to competition among bacteria as selective media inhibit other contaminating organisms.

*Coagulase-negative staphylococci (CONS)* were the most common bacteria isolated in this review, followed by gram-positive bacteria such as *Staphylococcus aureus*. However, *Staphylococcus aureus* is the most common bacterial infection in most countries and is responsible for over 1 million worldwide deaths, with no focus on global public health expenditure [10]. *Escherichia coli* were the most isolated gram-negative bacteria from mobile phones used by healthcare personnel. The possible reason for the high isolation of *Staphylococci* species might be related to their residence in mucosal systems and the isolation of *E. coli*, possibly due to cross-contamination with gastrointestinal samples. This finding was in line with findings from other studies [1, 2, 10, 16, 69].

The review’s second objective was to describe medication sensitivity and resistance patterns among African bacterial isolates. In a study conducted in Ethiopia, *ceftriaxone and ciprofloxacin* were effective against 71.7% and 89.1% of gram-positive bacterial isolates such as CONs and S. aureus, respectively, while *E. coli* was 100% sensitive to ciprofloxacin, gentamycin, and trimethoprim-sulfamethoxazole [42]. However, a study conducted in Nigeria found substantial resistance levels to *cotrimoxazole, tetracycline, ampicillin, gentamicin, ceftriaxone, and ciprofloxacin* [21]. Most patients who are treated at home are resistant to one or more antimicrobials [66]. Different bacterial strains, hospital environment, empirical treatment practice, use of antibacterial as a prophylactic, easy availability of some drugs without a prescription, drug dose, and indiscriminate/prolonged use of common antibiotics could all contribute to discrepancies in antimicrobial susceptibility in the included studies [70].

## Data Availability

The manuscript contains all pertinent information.

## Limitations

This research has certain limitations. Since all of the studies examined were cross-sectional studies in design, it could be difficult to establish a cause-effect relationship. The study’s findings were only generalizable to the included country’s sub-regions. In the end, only articles written in English were considered. Finally, future researchers should concentrate on observational studies with well-designed designs, such as cohort and interventional studies.

## Conclusion

The contamination of mobile phones used by HCWs with various bacterial isolates was found to be high in this review. The most common bacteria isolated were *coagulase-negative staphylococci, Staphylococcus aureus, and Escherichia coli*. The prevalence of bacterial contamination in mobile phones varies by country and sub-region. Healthcare workers should be required to practice proper hand hygiene and clean after using their phones in healthcare facilities. Thus, healthcare planners and policymakers should adopt guidelines to govern healthcare workers’ hand hygiene, disinfection, sterilization, and cleansing after using mobile phones in healthcare facilities.

## Ethics approval and consent to participate

Not applicable.

## Consent for publication

Not applicable.

## Availability of data and materials

The manuscript contains all pertinent information.

## Author’s contributions

DZ, BS, GB contributed to the conception, design, data extraction, and AM, FD, FN, DA, WN, MM, ZT and VC evaluated the methodological quality of the articles included, as well as the participated in data analysis, interpretation, writing the first draft of the paper. Finally, the authors read, commented on, edited, and approved the final draft version.

## Additional files

Additional file 1: PRISMA checklist (DOC)

Additional file 2: Search results of electronic databases (Screenshot)

Additional file 3: Risk of bias assessment of included studies (XLSX).

